# Health-related quality of life of patients presenting to the emergency department with a musculoskeletal disorder

**DOI:** 10.1101/2020.10.28.20221713

**Authors:** Rose Gagnon, Kadija Perreault, Jason Robert-Guertin, Simon Berthelot, Bertrand Achou, Luc J. Hébert

## Abstract

**Objectives:** The purpose of this study was to assess utility scores of patients presenting to the emergency department (ED) with a musculoskeletal disorder and to explore the influence of diverse factors on health-related quality of life.

**Design:** Secondary analysis of data obtained before randomization during a pragmatic randomized controlled trial

**Setting:** Academic ED in Quebec City (Canada)

**Participants:** Participants aged 18-80 years old presenting with a minor MSKD.

**Main Outcome Measures:** Health-related quality of life (five dimensions: mobility, self-care, usual activities, pain/discomfort, and anxiety/depression) and utility scores (0 – dead, 100 – perfect health), measured with the EQ-5D-5L, were compared between subgroups and with reference values using descriptive statistics (mean, median), rankFD ANOVAs, and χ^2^ tests.

**Results:** Sixty-nine participants completed the EQ-5D-5L. Mean and median utility scores were respectively 0.536 (95% CI: 0.479-0.594) and 0.531 (IQR: 0.356-0.760). Participants with higher levels of pain (<4/10: 0.741; 4-7/10: 0.572; >7/10: 0.433) or pain interference on function (<4/10: 0.685; 4-7/10: 0.463; >7/10: 0.294) presented significantly lower utility scores. No significant differences were found for other socio-demographic characteristics. The mean overall VAS score was 58.1 (95% CI: 52.2-64.0).

**Conclusions:** In patients with MSKDs presenting to the ED, higher levels of pain and pain interference on function may influence perceived health-related QoL. These findings need to be confirmed on a larger scale.

**Trial Registration:** This trial was registered at the US National Institutes of Health (ClinicalTrials.gov) #NCT04009369 on July 5, 2019

## Introduction

Musculoskeletal disorders (MSKDs) commonly refer to a group of symptoms and inflammatory or degenerative lesions of the musculoskeletal system in the neck, back, upper or lower limbs [1]. MSKDs are very common and are associated with various impairments such as pain, stiffness, loss of joint mobility and muscle strength, as well as bone deformity [2]. They are among the most disabling and costly non-fatal health conditions and their prevalence is expected to increase over the next decade [3–5]. If not managed promptly, they can have long-term consequences such as an increased pain level, loss of mobility and a greater risk of developing psychological symptoms [6, 7]. Patients presenting a MSKD that are not quickly managed also tend to make greater use of resources within the health care system, are more likely to be absent due to illness and experience greater loss of productivity at work [8–14]. Moreover, MSKDs can negatively affect patients’ daily activities and quality of life [6–8, 15, 16]. Many patients presenting a MSKD will consult the emergency department (ED) for their condition at some point due to a lack of adequate access to primary care services [17–19].

The EQ-5D-5L is a widely used preference-based multi-attribute health status classification system [20, 21]. It includes five dimensions representing different facets of daily life: mobility, self-care, usual activities, pain/discomfort, and anxiety/depression. These dimensions are scored on five levels ranging from no problem to unable/extreme problem. The EQ-5D-5L allows to derive a health state (i.e., 11111 to 55555) which can then be transformed into a measure of health-related quality of life (i.e., utility score). When compared to population norms, utility scores obtained in a subgroup with a condition such as a MSKD can provide a better understanding of the effects of a particular health condition on health related quality of life. A better understanding of the utility scores specific to certain disorders and the factors that may affect them could also allow better tailoring and prioritization of services and resources to these patients [22–24]. Although the EQ-5D-5L is one of the most widely used health-related quality of life measures to date, the majority of utility scores for patients with MSKDs have been published in Europe and very few have been published in North America. Moreover, to our knowledge, no utility scores have been published for MSKD patients in the ED, even though poor physical health-related quality of life is associated with an increased risk of ED visit [25].

Therefore, to address this lack of knowledge, the purpose of this study was to determine utility scores for patients presenting to the ED with a MSKD and to explore differences in health-related quality of life based on age, gender, triage score, annual income, self-reported other health conditions, onset of MSKD, presenting complaint, pain level and pain interference on function.

## Methods

### Study design and population

Data were acquired during a single center randomized controlled trial conducted between September 2018 and March 2019 in an Academic ED in Quebec City (Canada). In this trial, we recruited two groups of participants either having direct access to a physiotherapist in the ED or being managed according to usual practice by an emergency physician. The study was approved by the Research Ethics Committee of the CHU de Québec – Université Laval and registered at the US National Institutes of Health #NCT04009369. Patients presenting to the ED were recruited if they had a potential peripheral or vertebral minor MSKD and if given a triage score of 3 (urgent), 4 (less urgent) or 5 (non urgent) based on the Canadian Emergency Department Triage and Acuity Scale classification [26]. Other criteria for inclusion were being aged between 18 and 80 years old, having the ability to legally consent and participate, understanding French to complete the study questionnaires orally or in writing and being a beneficiary of the Quebec provincial health insurance plan (*Régie de l’assurance maladie du Québec*). The exclusion criteria were to present a major MSKD requiring emergent care (e.g. open fracture, dislocation, open wound), a red flag (e.g. progressive neurological disorder, infectious symptoms), a concomitant unstable clinical condition (e.g. pulmonary, cardiac, digestive and/or psychiatric) or being hospitalized or living in a long-term care facility.

### Study recruitment

Patients were considered for enrollment in the study based on information collected by the triage nurse and included within the electronic information system used at the ED to register patients. The research coordinator (RG) checked any new patient registered in the system to see if they were eligible and consented to participate in the study. Each participant was then asked to provide socio-demographic and clinical data and complete baseline data on pain intensity, pain interference with function and quality of life prior to their consultation with the emergency physician or physiotherapist.

### Measures

Our primary outcome was the health-related quality of life at the ED visit measured using the EQ-5D-5L descriptive system [27]. The EQ visual analog scale (VAS) ranging from 0 (worst imaginable health state) to 100 (best imaginable health state) was also used. The EQ-5D-5L has been recognized as being reliable, valid and responsive [28, 29]. The questionnaire was administered using paper format in the ED. Health utility scores were calculated using the Canadian value set developed by Xie et al. using a composite time trade-off (traditional and lead time trade-off) [20]. Utility scores based on this value set range from −0.148 (55555 - worst health state) to 0.949 (11111 - best health state) depending on the health state reported by the participant with the EQ-5D-5L.

Several demographic and clinical characteristics that might affect health-related quality of life, such as age, gender, triage score, annual income, self-reported other health conditions, onset and region of the presenting complaint were compiled using the initial questionnaire, as mentioned above. Pain intensity was measured using the *Numeric Pain Rating Scale*, an 11-point scale ranging from 0 (no pain) to 10 (worst pain imaginable). Pain interference with function was measured using the Pain Inventory subscale of the short version of the *Brief Pain Inventory*. This subscale covers ten activities of daily living: general activity, mood, walking, work, sleep, personal care, etc. For each, the respondent is asked to indicate the extent to which pain experienced in the past 7 days interfered with each of the activities on a scale of 0 to 10, where 10 means that the pain experienced completely interfered with the activity and 0 that pain doesn’t interfere with activity. Both tools have been shown to be reliable, valid and responsive [30–34].

### Data analysis and sample size

Socio-demographic, utility and VAS scores were calculated using descriptive statistics (both mean and median scores were produced for utility and VAS scores). We used non-parametric repeated-measures analyses of variance (one-way and two-way rankFD ANOVAs) to compare median utility scores in the different subgroups (R software, 4.0.2; package rankFD, 0.0.5; proc rankFD). χ^2^ tests were used to compare the distribution of problems reported by dimension and demographic characteristics (SPSS software, version 25; IBM, Armonk, NY USA). The α criterion was set at 0.05 for all statistical analyses.

## Results

### Participants

The eligibility of 589 patients was assessed and 78 were recruited (Fig. 1). The EQ-5D-5L was completed by 69 of the 78 (88.5%) participants recruited (Fig. 1). In terms of baseline characteristics, the majority of participants recruited were between 18 and 65 years of age, male, and had an income level considered low to moderate (Table 1). Most participants did not report any associated health condition, had a triage score of 4 (less urgent) and presented a complaint related to the spine or the lower limb. For more than 80% of all participants, the onset of their condition occurred within the last two weeks (Table 1). Nearly 90% of the participants had a pain level greater than 4/10 and 50% of the participants had pain interference with function between 4 and 7/10.

**Table 1.** Baseline characteristics of study participants (n=69)

| Characteristics | Sample (%) | 95% CI (%) |
| --- | --- | --- |
| Age, yrs | n=69 |  |
| 18-65 | 88.4 | 77.9 – 94.5 |
| > 65 | 11.6 | 5.5 – 22.1 |
| Gender | n=69 |  |
| Women | 42.0 | 30.4 – 54.5 |
| Men | 58.0 | 45.5 – 69.6 |
| Triage score | n=69 |  |
| Urgent (P3) | 42.6 | 30.4 – 54.5 |
| Semi-Urgent (P4) | 57.4 | 44.1 – 68.2 |
| Gross income, \$ CAN | n=63 | |
| 1 – 49 999 | 65.1 | 51.9 – 76.4 |
| 50 000 – 99 999 | 34.9 | 23.6 – 48.1 |
| Self-reported other health conditions | n=69 |  |
| No | 58.0 | 45.5 – 69.6 |
| Yes | 42.0 | 30.4 – 54.5 |
| Onset of, weeks | n=67 |  |
| 0-2 | 82.1 | 70.4 – 90.0 |
| > 3 | 17.9 | 10.0 – 29.6 |
| Presenting complaint | n=67 |  |
| Upper limb | 11.9 | 5.7 – 22.7 |
| Lower limb | 38.8 | 27.4 – 51.5 |
| Cervical spine | 22.4 | 13.5 – 34.5 |
| Thoracic and lumbar spine | 26.9 | 17.1 – 39.3 |
| Pain level, /10 | n=68 |  |
| < 4 | 11.2 | 5.6 – 22.4 |
| 4 to 7 | 48.5 | 36.4 – 60.9 |
| > 7 | 39.7 | 28.3 – 52.3 |
| Pain interference, /10 | n=69 |  |
| < 4 | 40.6 | 29.1 – 53.1 |
| 4 to 7 | 50.1 | 38.5 – 62.9 |
| > 7 | 8.7 | 3.6 – 18.6 |

**Fig. 1.**
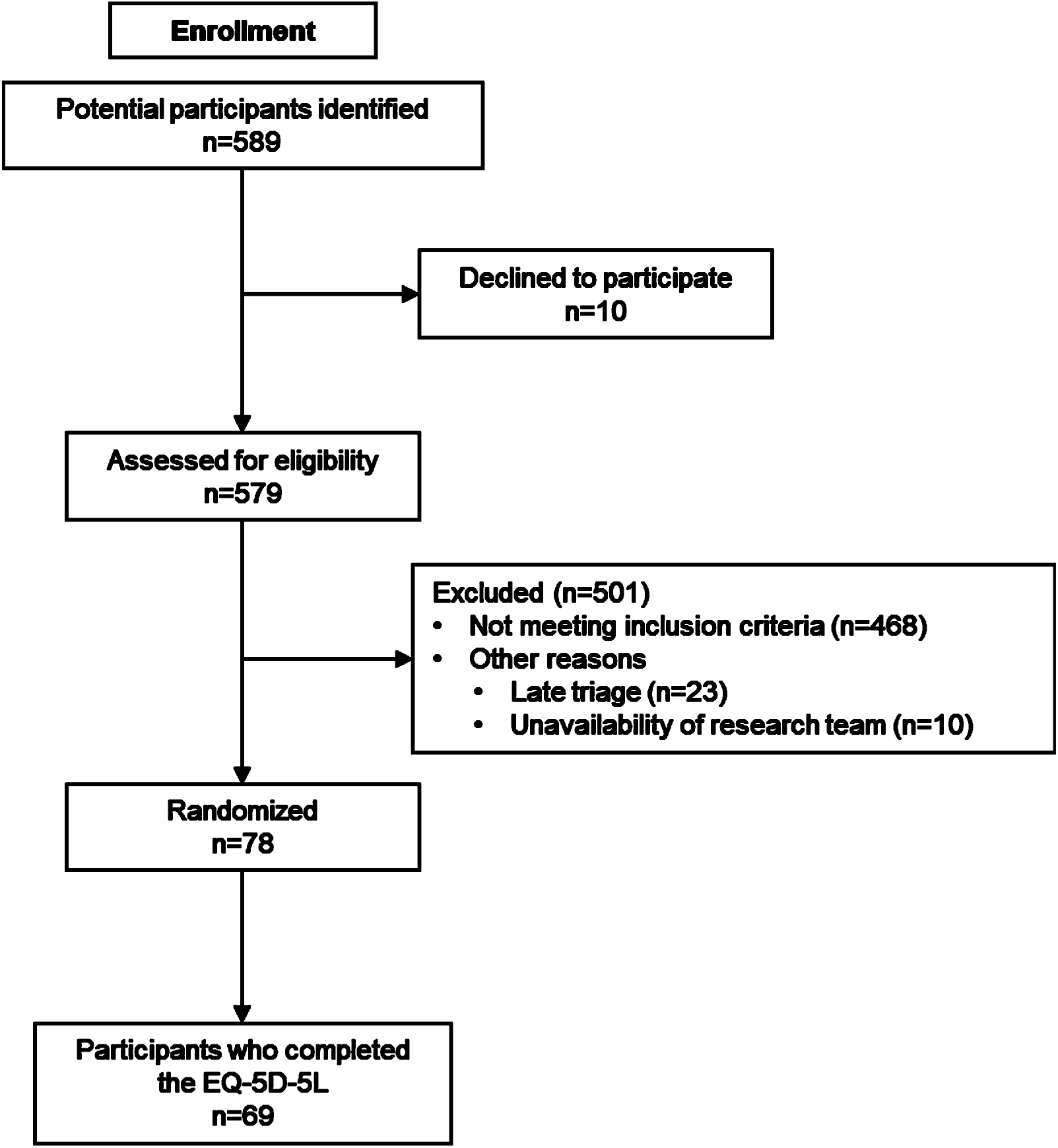
CONSORT Flow Diagram of participants’ recruitment.

### Overall EQ-5D-5L utility scores and EQ-VAS scores

Table 2 lists all mean and median utility scores according to the demographic and clinical characteristics presented earlier. Mean and median overall health-related utility scores were respectively 0.536 (95% CI: 0.479-0.616) and 0.540 (IQR: 0.357-0.782). The mean overall EQ-VAS score was 58.1 (95% CI: 52.2-64.0). The lowest utility scores were between 0.00 and 0.10 and the highest ones, between 0.90 and 0.949 (Fig. 2). Forty-seven percent of participants had a utility score comprised between 0.00 and 0.50 (Fig 2.). EQ-VAS scores ranged from 5 to 100 and more than 30.9% of participants presented a score between 5 and 50 (Fig. 3).

**Table 2.** EQ-5D-5L utility scores and EQ-VAS scores (n=69)

|  | EQ-5D-5L |  |  |  |  |
| --- | --- | --- | --- | --- | --- |
|  | Mean | (95% CI) | Median | (IQR) | p-value |
| Overall | 0.536 | (0.479-0.594) | 0.531 | (0.356-0.760) |  |
| EQ-VAS | 58.1 | (52.2-64.0) | 60.0 | (40.0-60.0) |  |
| Age, years |  |  |  |  | .129 |
| 18-65 | 0.554 | (0.492-0.616) | 0.540 | (0.357-0.782) |  |
| > 65 | 0.412 | (0.253-0.571) | 0.443 | (0.219-0.574) |  |
| Gender |  |  |  |  | .780 |
| Women | 0.541 | (0.449-0.635) | 0.564 | (0.372-0.730) |  |
| Men | 0.532 | (0.457-0.608) | 0.530 | (0.346-0.779) |  |
| Triage score |  |  |  |  | .222 |
| Urgent (P3) | 0.489 | (0.402-0.577) | 0.466 | (0.314-0.660) |  |
| Semi-Urgent (P4) | 0.561 | (0.483-0.639) | 0.558 | (0.357-0.783) |  |
| Gross income, \$ CAN | | | | | .225 |
| 1 – 49 999 | 0.578 | (0.504-0.652) | 0.606 | (0.437-0.782) |  |
| 50 000 – 99 999 | 0.503 | (0.391-0.616) | 0.452 | (0.317-0.769) |  |
| Self-reported other health conditions |  |  |  |  | .209 |
| No | 0.570 | (0.489-0.650) | 0.616 | (0.357-0.783) |  |
| Yes | 0.492 | (0.409-0.575) | 0.474 | (0.357-0.622) |  |
| Onset of, weeks |  |  |  |  | .377 |
| 0-2 | 0.538 | (0.473-0.604) | 0.531 | (0.366-0.776) |  |
| > 3 | 0.472 | (0.342-0.603) | 0.462 | (0.254-0.637) |  |
| Presenting complaint |  |  |  |  | .239 |
| Upper limb | 0.626 | (0.467-0.784) | 0.623 | (0.455-0.784) |  |
| Lower limb | 0.470 | (0.388-0.553) | 0.437 | (0.328-0.611) |  |
| Cervical spine | 0.602 | (0.469-0.736) | 0.656 | (0.426-0.743) |  |
| Thoracic and lumbar spine | 0.496 | (0.354-0.638) | 0.477 | (0.254-0.763) |  |
| Pain level, /10 |  |  |  |  | .019 |
| < 4 | 0.741 | (0.501-0.980) | 0.867 | (0.610-0.882) |  |
| 4 to 7 | 0.572 | (0.500-0.644) | 0.616 | (0.402-0.744) |  |
| > 7 | 0.433 | (0.347-0.518) | 0.437 | (0.257-0.549) |  |
| Pain interference, /10 |  |  |  |  | .001 |
| < 4 | 0.685 | (0.605-0.764) | 0.753 | (0.532-0.862) |  |
| 4 to 7 | 0.463 | (0.394-0.533) | 0.450 | (0.336-0.612) |  |
| > 7 | 0.294 | (0.126-0.463) | 0.321 | (0.119-0.431) |  |

CI: confidence interval, IQR: interquartile range

**Fig. 2.**
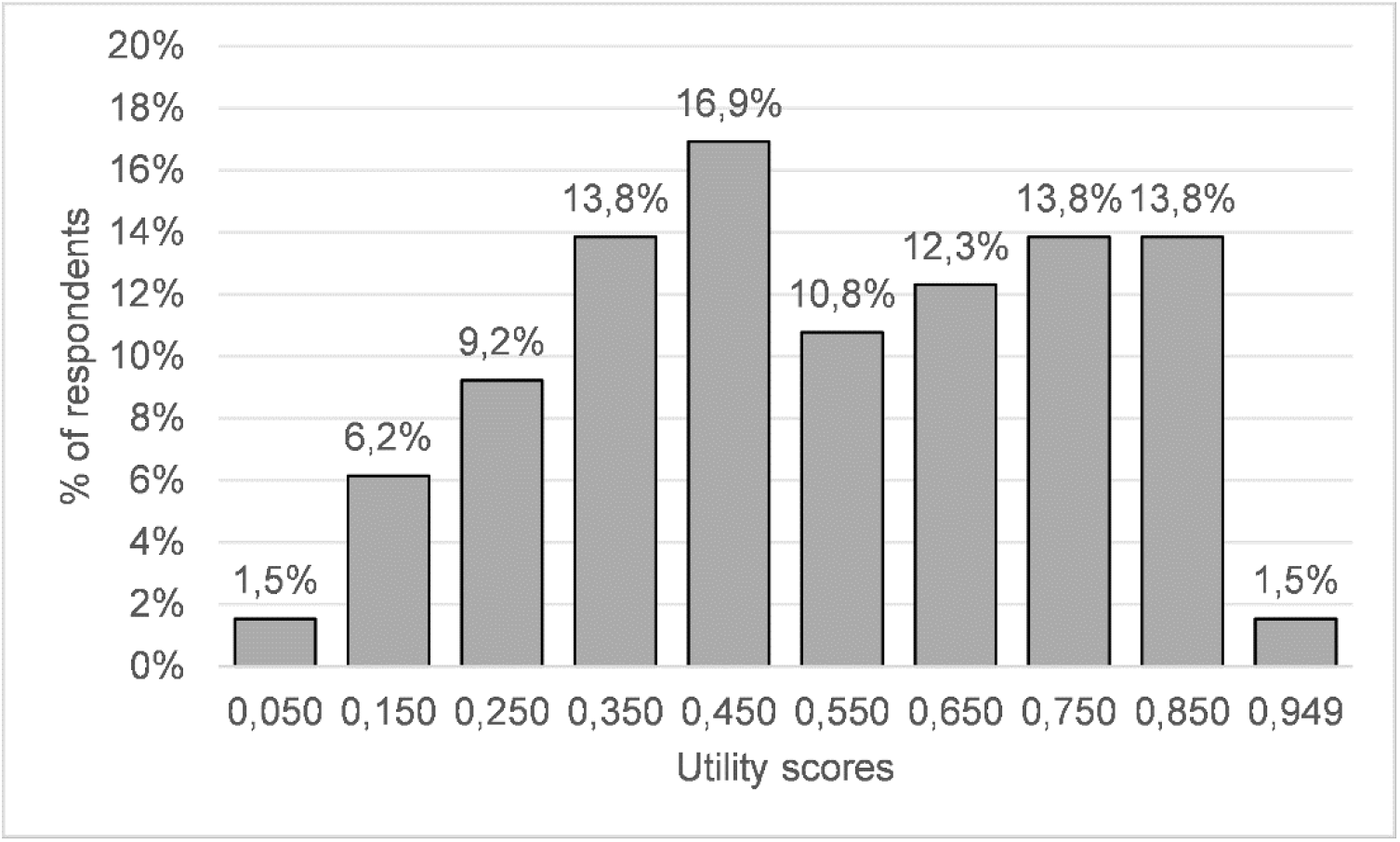
Utility scores distribution for EQ-5D-5L. The utility scores on the x-axis represent the central value of each band. The first band is from 0.000 to 0.100 (0.050 for the central value) and the last band corresponds to the ceiling effect (0.949).

**Fig. 3.**
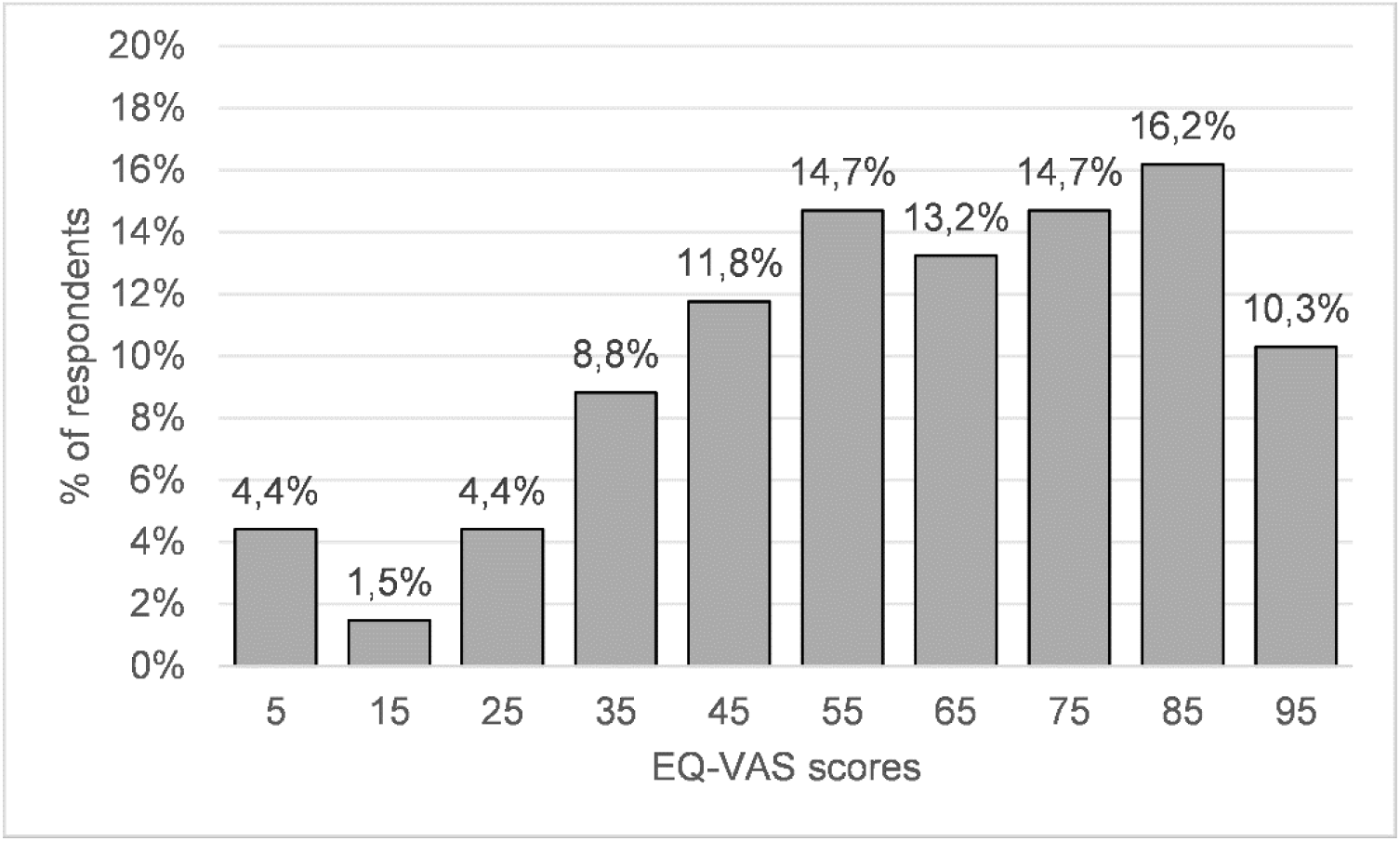
EQ-VAS scores distribution. The EQ-VAS scores on the x-axis represent the central value of each band. The first band is from 0 to 10 (5 for the central value) and the last band is from 90 to 100 (95 for the central value).

### Differences in scores according to demographic and clinical characteristics

No significant differences were found in subgroup scores as for age, gender, triage score, annual income, presence of a self-reported other health condition, onset and region of the presenting complaint. Participants who reported a higher level of pain during their ED visit had significantly lower utility scores than those with a lower pain level (p=.019, Table 2). A higher level of pain interference on function was also associated with a significantly lower utility score (p=.001).

When considering the five dimensions of the EQ-5D-5L separately, 72.5% of participants considered having mobility problems, 58.0% self-care problems, 89.7% problems with their usual activities, 97.0% pain or discomfort and 46.4% anxiety or depression (Table 3). No significant differences were found between women and men or between participants with a triage score of 3 or 4 in the distribution of problems reported (Table 3). However, women reported less pain/discomfort (Pain severe to extreme, Men: 52.6%, Women: 42.9%) and more anxiety/depression (Presence of anxiety/depression, Men: 42.5%, Women: 51.7%) than men (Table 3). Participants with a triage score of 3 reported more problems with their usual activities (Severe problems to unable, 3: 50.0%, 4: 28.2%) and more anxiety or depression (Presence of anxiety/depression, 3: 51.7%, 4: 43.6%) (Table 3).

**Table 3.** Distribution of problems reported by dimension and demographic characteristics.

|  | Total<br>n=69 | Women<br>n=29 | Men<br>n=40 | p-value | Triage<br>score 3<br>n=29 | Triage<br>score 4<br>n=39 | p-value |
| --- | --- | --- | --- | --- | --- | --- | --- |
| **Mobility, %** |  |  |  | .271 |  |  | .086 |
| No problem | 27.5 | 27.6 | 27.5 |  | 13.8 | 35.9 |  |
| Slight problems | 13.0 | 10.3 | 15.0 |  | 24.1 | 5.1 |  |
| Moderate problems | 21.7 | 31.0 | 15.0 |  | 27.6 | 17.9 |  |
| Severe problems | 24.6 | 13.8 | 32.5 |  | 17.2 | 30.8 |  |
| Unable | 13.0 | 17.2 | 10.0 |  | 17.2 | 10.3 |  |
| **Self-care, %** |  |  |  | .604 |  |  | .266 |
| No problem | 42.0 | 41.4 | 42.5 |  | 27.6 | 51.3 |  |
| Slight problems | 20.3 | 13.8 | 25.0 |  | 31.0 | 12.8 |  |
| Moderate problems | 29.0 | 34.5 | 25.0 |  | 27.6 | 30.8 |  |
| Severe problems | 7.2 | 6.9 | 7.5 |  | 10.3 | 5.1 |  |
| Unable | 1.4 | 3.4 | 0.0 |  | 3.4 | 0.0 |  |
| **Usual Activities, %** | n=68 |  | n=39 | .327 | n=28 |  | .418 |
| No problem | 10.3 | 13.8 | 7.7 |  | 7.1 | 12.8 |  |
| Slight problems | 19.1 | 27.6 | 12.8 |  | 14.3 | 20.5 |  |
| Moderate problems | 33.8 | 31.0 | 35.9 |  | 28.6 | 38.5 |  |
| Severe problems | 27.9 | 17.2 | 35.9 |  | 35.7 | 23.1 |  |
| Unable | 8.8 | 10.3 | 7.7 |  | 14.3 | 5.1 |  |
| **Pain/Discomfort, %** | n=66 | n=28 | n=38 | .600 | n=28 | n=37 | .398 |
| No | 3.0 | 7.1 | 0.0 |  | 3.6 | 2.7 |  |
| Slight | 13.6 | 14.3 | 13.2 |  | 10.7 | 13.5 |  |
| Moderate | 34.8 | 35.7 | 34.2 |  | 32.1 | 37.8 |  |
| Severe | 45.5 | 39.3 | 50.0 |  | 50.0 | 43.2 |  |
| Extreme | 3.0 | 3.6 | 2.6 |  | 3.6 | 2.7 |  |
| **Anxiety/Depression, %** |  |  |  | .834 |  |  | .960 |
| No | 53.6 | 48.3 | 57.5 |  | 48.3 | 56.4 |  |
| Slight | 27.5 | 27.6 | 27.5 |  | 31.0 | 25.6 |  |
| Moderate | 14.5 | 17.2 | 12.5 |  | 17.2 | 12.8 |  |
| Severe | 2.9 | 3.4 | 2.5 |  | 3.4 | 2.6 |  |
| Extreme | 1.4 | 3.4 | 0.0 |  | 0.0 | 2.6 |  |

## Discussion

The aim of our study was to determine utility scores for patients presenting to the ED with a MSKD and to explore differences in health-related quality of life based on age, gender, triage score, annual income, self-reported other health conditions, onset of MSKD, presenting complaint, pain level and pain interference on function. Participants with higher pain level or pain interference on function reported significantly lower health-related quality of life. No significant differences were found for all other socio-demographic characteristics.

To our knowledge, no EQ-5D-5L utility scores have been published for MSKD patients presenting to the ED. However, Quebec population reference values were published in 2019 by Poder et al [35]. In their study, the overall mean health related utility score was 0.824 (95% CI: 0.818-0.829). Using the EQ-5D-5L minimally important difference (MID: 0.074) as a comparison, this score is clinically higher than the value obtained in our study at 0.536 (95% CI: 0.479-0.594) [36]. Also, according to Poder et al., in patients with MSKDs other than osteoarthritis, the mean utility score was 0.586 (95% CI: 0.546-0.627), which is higher than the score obtained in our study, but not clinically different. Poder et al. report mean utility score in people experiencing pain of 0.631 (95% CI: 0.607-0.654) for all persons presenting pain (all pain levels) in Quebec’s population who responded to the EQ-5D-5L (n=367). Although this value seems at first glance to be lower than the one obtained in participants of our study with a pain level of less than 4/10 (0.741, 95%: 0.501-0.980), it is important to remember that we may still have a clinically lower mean utility score if we had aggregated our three pain categories (< 4, 4 to 7, > 7). The low scores could be partially explained by the fact that patients consulting the ED for a MSKD are often more concerned about their condition. In fact, certain factors specific to patients, such as having a chronic disease, having already experienced a serious health problem or having the perception of being in poorer health, would also influence the use of the ED for patients with MSKDs [17]. However, our sample included fewer women and had a lower average income level than those of the population of Quebec. Nevertheless, the two samples were similar in terms of distribution of the age groups surveyed.

In our study, health-related quality of life appeared to be influenced only by higher levels of pain and pain interference on function. These results somewhat contradict those in the literature, in that some studies have shown that socio-demographic characteristics such as age, gender, and income level might influence health-related quality of life [35, 37]. However, these results are from healthy participants, not from people presenting with a MSKD. Indeed, although MSKDs affect 20 to 33% of the world’s population and cost the healthcare system several billion dollars annually [38], very few studies have looked at the factors influencing health-related quality of life in this sub-population specifically. Those that have done so often focus on a specific disorder (e.g. arthritis, scoliosis, total joint replacement, etc.), and not on a sample with a set of disorders representative of this population [39–42]. Considering the very high prevalence of MSKDs and the expected increase in the number of seniors in the next decade [3], a better understanding of factors impacting on health-related quality of life would help better manage patients with MSKDs and ED managers to better tailor ED services and resources to these patients.

## Limitations

Our study is the first to report utility scores in a sample of patients presenting to the ED for a MSKD. Another strength of our study is that we analyzed the influence of various socio-demographic factors on health-related quality of life.

Nevertheless, our results should be interpreted with caution. Our sample size was small, so some of the subcategories presented have relatively few participants. Studies with larger sample size will be required to confirm our results. Finally, our sample should have contained more participants aged 65 and over to be more representative of the general population and to allow for more in-depth analyses of this subgroup of interest. Still, several studies conducted in the ED among people with MSKDs report average ages similar to our own [15, 43–45]. It is therefore reasonable to believe that our sample remains representative of the population consulting the ED for a MSKD.

## Conclusion

In MSKD patients presenting to the ED, health-related utility scores are below the Quebec population reference values and higher levels of pain and pain interference on function may influence their perceived health related quality of life. These findings need to be confirmed using multicentre studies with a larger sample size.

## Data Availability

The data that support the findings of this study are available from the corresponding author, LJH, upon reasonable request.

## Acknowledgements

We would like to thank the following persons for their contribution to project implementation : All project participants, Antony Barabé PT, physiotherapist at the Centre Hospitalier de l’Université Laval (CHUL), the entire team of managers at the Direction des services multidisciplinaires of the CHU de Québec – Université Laval (Marie-Christine Laroche, Catherine van Neste, Marie-Claude Brodeur and Stéphane Tremblay) for their support throughout the implementation of the project and its realization, and Mathieu Blanchet, MD, FRCPC, head of the CHUL ED department during the duration of the study.

## Declarations

### Ethics approval

This trial was approved by the Research Ethics Committee of the CHU de Québec - Université Laval #MP-20-2019-4307

### Consent to participate

Informed consent was obtained from all individual participants included in the study.

### Authors contributions

RG, KP, SB, BA and LJH were involved in study concept and design. RG, KP, SB and LJH were involved in acquisition of the data. RG, KP, JRG, BA and LJH were involved in analysis and interpretation of the data. RG drafted the manuscript. KP, JRG, SB, BA and LJH were involved in critical revision of the manuscript for important intellectual content. RG, KP and LJH were involved in acquisition of funding.

